# Healthcare Worker Staffing Ratios Affect Methicillin-Resistant *Staphylococcus aureus* Acquisition

**DOI:** 10.1101/2024.02.14.24302485

**Authors:** Stephanie Sikavitsas Johnson, Matthew Steven Mietchen, Eric Thomas Lofgren

## Abstract

**Importance:** This study addresses the pressing clinical question of how variations in physician and nursing staffing levels influence methicillin-resistant *Staphylococcus aureus* (MRSA) rates, providing essential insights for optimizing staff allocation and improving patient outcomes in critical care settings.

**Objective:** The main objective is to assess whether variations in staffing ratios and workload conceptualization significantly alter the rates of MRSA acquisitions in the ICU setting.

**Design:** This simulation-based study utilizes stochastic compartmental mathematical modeling to explore the impact of staffing ratios and workload conceptualization on MRSA acquisitions in ICUs. Derived from a previously published model, the analysis involves running year-long stochastic simulations for each scenario 1000 times, varying nurse-to-patient ratios and intensivist staffing levels under infinite and finite workload conceptualizations. Our baseline model was a 3:1 nurse ratio with one intensivist.

**Main Outcome:** MRSA acquisitions in ICUs, measured as median acquisitions per 1000 person-years.

**Results:** Under baseline conditions, our model had a median of 8.2 MRSA acquisitions per 1000 person-years. Varying patient-to-nurse ratios and intensivist numbers showed substantial impacts. For infinite models, a 2:1 nurse ratio resulted in a 21% decrease, while a 1:1 nurse ratio led to a 65% reduction. Finite models demonstrated even larger effects, with a 48% decrease when having a 2:1 ratio, and an 83% reduction with a 1:1 nurse ratio. Reducing patient-to-nurse ratios in finite models increased acquisitions exponentially with a 348% increase for a 6:1 ratio. Intensivist variations had modest impacts.

**Conclusions and Relevance:** Our study highlights the crucial role of optimizing staffing levels in ICUs for effective MRSA infection control. While intensivist variations have modest effects, bolstering nursing ratios significantly reduces MRSA acquisitions, underscoring the need for tailored staffing strategies, and recognizing the nuanced impact of workload conceptualization. Our findings offer practical insights for refining staffing protocols, emphasizing the dynamic nature of healthcare-associated infection outcomes.

**Key Points:** *Question:* How does the conceptualization of ICU healthcare worker tasks in models—whether infinite or finite— impact the results of changes in staffing ratios affecting methicillin-resistant *Staphylococcus aureus* (MRSA) acquisition?

*Findings:* In this compartmental mathematical model approach that included 15 different models, the trends of the impact of staffing ratios were consistent between the Infinite and Finite tasks models. However, both the absolute and relative values were markedly different, with the infinite task models having a much more linear effect on MRSA acquisitions while the number of MRSA cases in the finite model continued to rise exponentially as the number of nurses decreased.

*Meaning:* It is essential when considering model generalizability, to state the assumptions made about how workload and contact patterns within a hospital work, and to ensure these are appropriately tailored for the specific setting being modeled.

## Background

Infection prevention staff deliver infection control and effective care at the intersection of patient care, workforce compliance, and hospital budgets. One of the largest challenges affected is staffing ratios, an issue that not only impacts the quality of patient care but also plays a pivotal role in safeguarding patients from HAIs^1–3^. Staffing issues have been shown to impact many things e.g. fatigue, mortality, and infection control^1,2,4^. Only one state, California, has a mandated ICU patient-to-nurse ratio of 2:1 ^5^. Other states have left this up to hospitals and healthcare systems, leading to a national average of 3:1 that is used as a baseline for many modeling studies ^2,6–8^.

Hospital modeling provides a unique vantage point for understanding the dynamics within specific hospital units-such as intensive care units (ICUs). These models can help critical care teams and infection control professionals understand the intricate interplay between their own ward’s behaviors and healthcare-associated infections (HAIs)^9–12^. Modeling studies have emerged as indispensable tools, especially in scenarios or with pathogens where conducting randomized controlled trials would be cost-prohibitive, labor-intensive or the pathogens cause sporadic and self-limiting epidemics^7,9,10,13–15^.

Methicillin-resistant *Staphylococcus aureus* (MRSA) is found in approximately 9% of ICU admissions, is environmentally shed, transmitted through contact, and is responsible for sporadic outbreaks and is a priority for infection control due to their antibiotic-resistance. These characteristics render it an excellent candidate for modeling burden-mitigating strategies in infection control^10–12,16^.

While studies have shown that lower staffing is good for patients, less is known about how the mix of physician and nursing staffing affects prevention efforts^1,3,17^. Does adding another physician make the other physician’s workload less but add more patient contact as a whole – and what are the implications of that for infection transmission? If a unit has to increase their staffing ratio, does that mean an increased workload for the remaining nurses, or are they already at capacity and those tasks just get added in, meaning less care and contact with nurses? These scenarios happen frequently, and must be taken into consideration at both the hospital and unit level.^10^.

This study focuses on understanding how one structures a model affects the results a healthcare team might be expected to see. This was done by modifying a previously published model of an ICU, and adjusting the staffing ratios of nurses and physicians to obtain the number of yearly MRSA acquisitions based on staffing^7^. In addition, we adjusted the models to have nurses and physicians have varying amount on contact with patients. MRSA was used because it is a well-studied pathogen in healthcare infection that is primarily spread via human interaction in hospital settings ^3,11,18–20^. There are over 300,00 hospitalized cases a year, with an estimated 11,00 deaths and hospital-onset MRSA bloodstream infections have been stable since 2013^20^.

## Methods

### Model Structure

Our baseline model of MRSA transmission is based on a previously published model of a 18-bed medical ICU, staffed by six nurses (a 3:1 patient-to-nurse ratio) and one dedicated intensivist (see Figure 1) ^4,6,7^. The 18-bed size was chosen for coherence with previous studies and for ease of creating a variety of whole numbered patient: staff ratios^4,17^. Our parameters came from previously published models of ICU-based MRSA transmission and are shown in Table 1 ^6,7,16,21^.

**Table 1.**
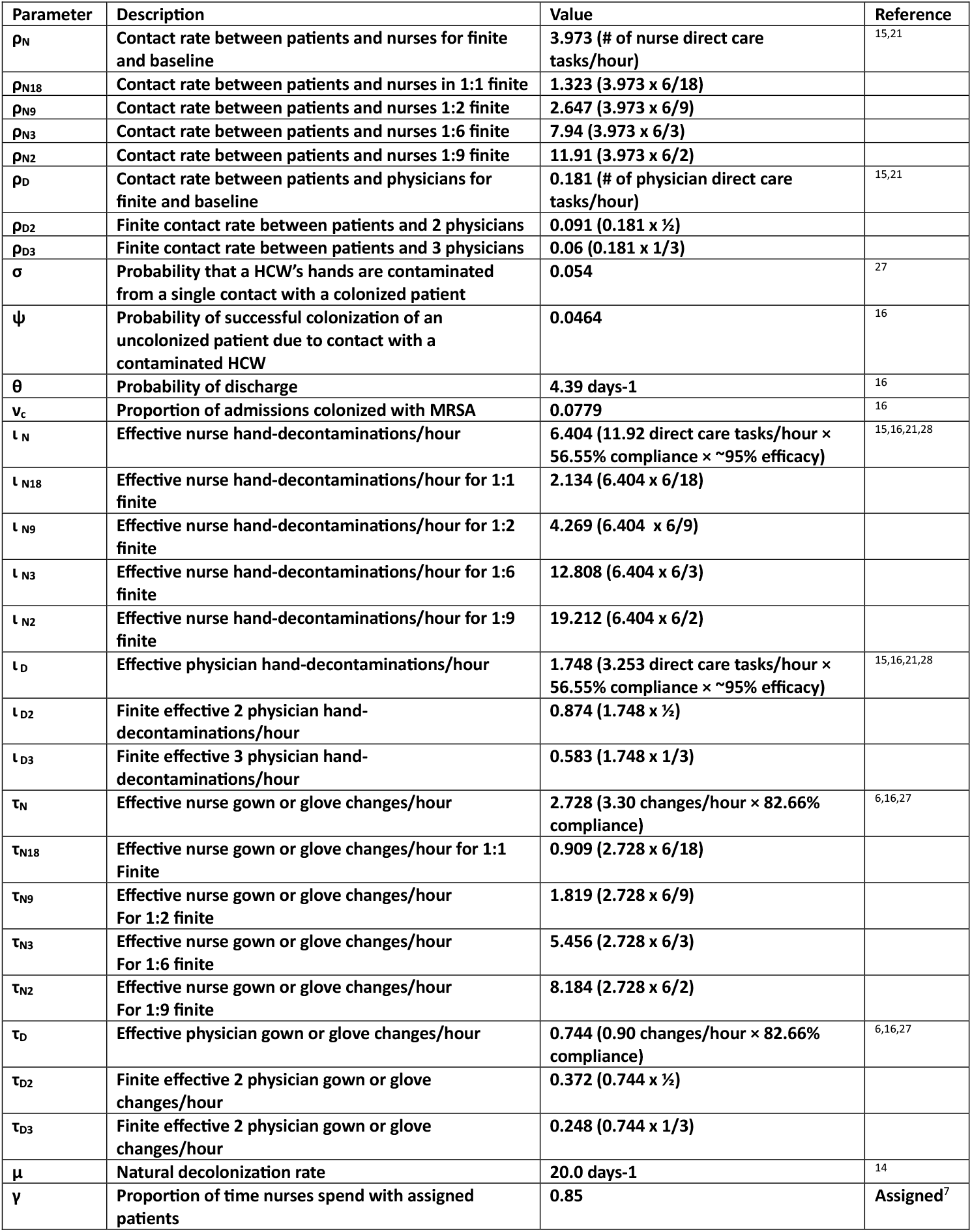
Parameters for Modeling the Effect of Staffing Levels on Methicillin-Resistant *Staphylococcus aureus* Acquisitions in an ICU.

**Figure 1.**
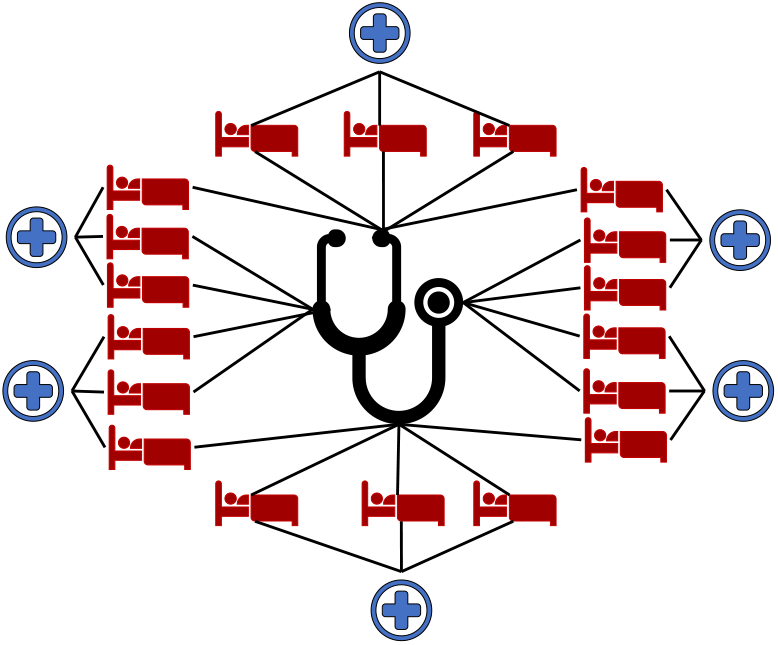
Baseline metapopulation compartmental model of methicillin-resistant *Staphylococcus aureus* (MRSA) acquisition on varying staffing levels and the multiple staffing level models.

Each nurse was assigned a specific cohort of three patients and were with them 85% of the time, but also interacted with patients outside their cohort due to a number of reasons (e.g., staff breaks, patient care needing more than one nurse, cross-coverage), expressed as the parameter gamma, (γ) (Table 1). Both within and outside each assigned cohort, the nurses are assumed to see their patients randomly. Intensivists see all patients in the ICU and further addition of intensivists does not result in cohorting.

Healthcare workers are represented by two possible states, either uncontaminated (D_u_ or N_u_) or contaminated (D_c_ or N_c_), and patients are either uncolonized or colonized (P_u_ or P_c_). Doctors and nurses have separate contact rates with patients, with doctors having less direct care tasks (touching patients or the immediate surrounding environment) as well as hand hygiene and gowning/gloving opportunities than nurses. The model equations are available in Supplemental Table 1.

This model makes a number of simplifying assumptions. First, patients are confined to their single-bed occupancy rooms and do not interact with anyone other than the nurses and doctors. We assume that nurses and doctors can only contaminate patients and vice versa, and do not transmit MRSA to other healthcare workers via direct contact or environmental exposure. Our model has the ICU as a “closed ICU” so that only people working in the ICU are allowed to interact with the patients. The ICU is also considered to be always at 100% capacity, so if a patient leaves, another one is admitted automatically^22^. We assume that hand hygiene and donning/doffing, while done when SHEA guidelines recommend, are done with imperfect compliance^23^. Finally, we assume that MRSA colonization is detected instantly with perfect sensitivity and specificity, but that there are no existing treatment or decolonization procedures being performed, save for a low baseline rate of natural decolonization (μ).

### Staffing Simulations

Fifteen staffing scenarios were implemented, ranging from a 9:1 patient: nurse ratio to a 1:1 patient: nurse ratio, as well as varying the number of intensivists in the ICU from 1 to 3.

We also consider two different paradigms to conceptualize how work within an ICU is performed, and how staffing levels might impact that work. The first, which we term the “Infinite Task Model”, posits that there are effectively infinitely many tasks to perform within an ICU in a given shift. As a result, the addition of new staff increases the *number* of tasks performed, but the *per-staff* contact rate between patients and staff members remains constant but potentially changing the overall contact rate between patients and staff – effectively, the ICU staff as a whole may now accomplish more, but individual-level workloads are not appreciably decreased. The second, termed the “Finite Task Model”, posits a large but ultimately fixed number of tasks to be accomplished in a given shift. Here, the addition of a new staff member proportionately decreases the number of tasks each healthcare worker has to do, altering the per-staff contact rate, but keeping the overall contact rate fixed.

Each of the thirty possible scenarios (five levels of nurse staffing by three levels of intensivist staffing for both infinite and finite models) was simulated stochastically 1000 times by means of Gillespie’s Direct Method to obtain a distribution of the number of MRSA acquisitions over a year, which was used as our primary outcome. Due to the non-normalcy of the resulting distributions, differences between staffing levels were analyzed with nonparametric Kruskal-Wallis tests. The models were written and simulated with Python 3.6 using the StochPy package^24^. Statistical analysis and visualization was done in R v4.2.2. A formal description of the model using the MInD Framework^25^ may be found in the Supplemental Information and code and model output are available at https://github.com/epimodels/StaffRatios.

## Results

### Baseline

Our baseline model had a median of 8.2 acquisitions per 1000 person-years (IQR 7.2-9.4).

### Infinite Task Models

Using the infinite tasks model, the average number of MRSA acquisitions was significantly different between the baseline and the adjusted nursing ratios. Going to a 2:1 patient: nurse ratio had a 21% decrease of MRSA acquisitions (χ2 =500, p-value <0.05), having a 1:1 patient: nurse ratio decreased it 65% (χ2 =1409, p-value <0.05). Meanwhile, going from a 3:1 ratio to a 6:1 nurse ratio increased acquisitions 37% (Figure 2A). Similar patterns were observed with the two doctor and three doctor models (Figure 2B-C).

**Figure 2.**
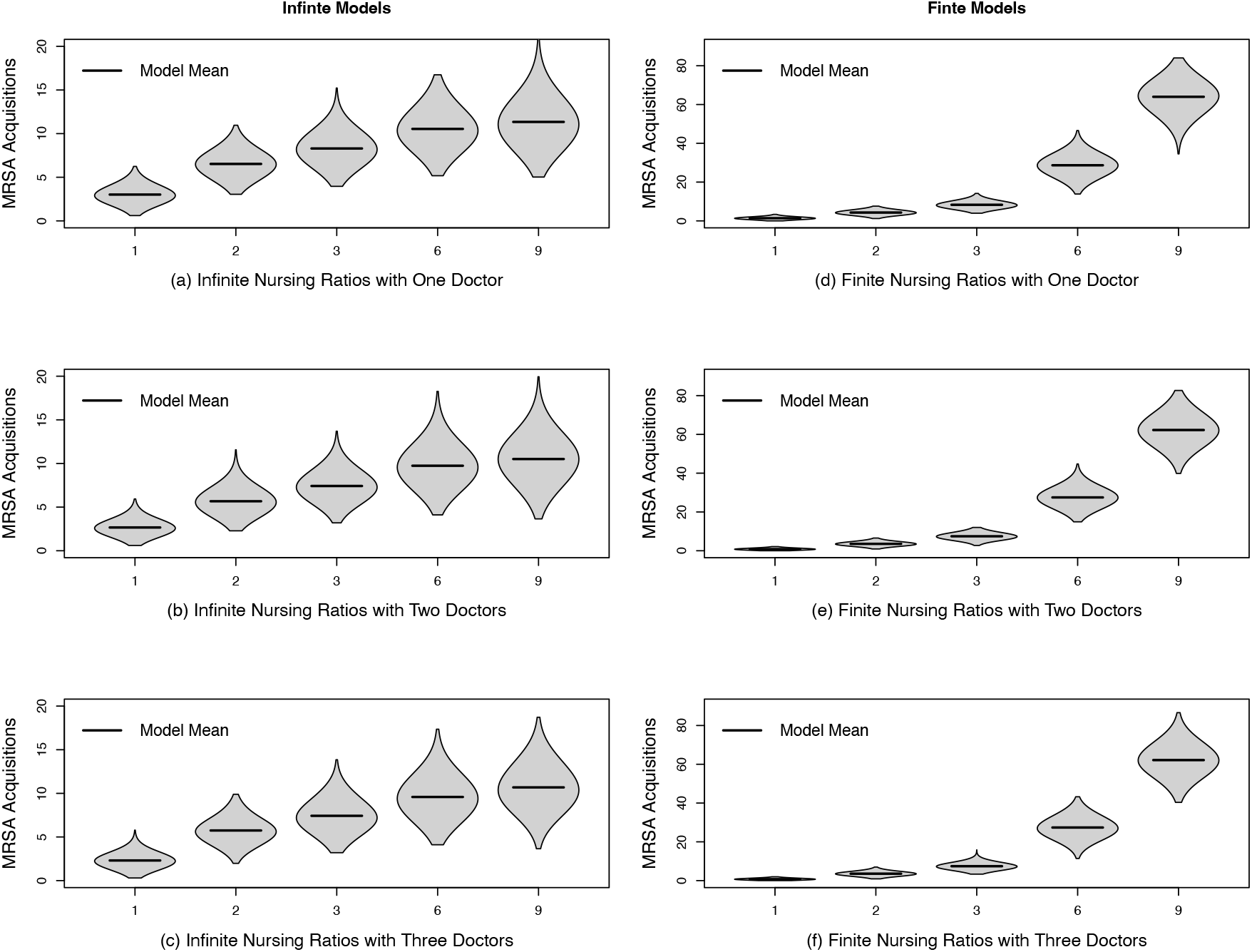
Distribution of cumulative MRSA acquisitions per 1000 patient-days in 1,000 simulated 18-bed ICUs under the different staffing ratios with both Finite and Infinite Task Scenarios.

### Finite Task Models

Using the finite tasks model, there was a significant difference between the baseline and all other modeled scenarios. Going to a 2:1 nurse ratio decreased acquisitions by 48% (χ2 =1398, p-value <0.05), or 83% reduction if there was a 1:1 nurse ratio (χ2 =1501, p-value <0.05) (Figure 2D). Reducing nurses had a much larger impact as well, a 6:1 ratio increased acquisitions by 348%, or 779% if the ICU was at a 9:1 nurse ratio. Once again this was observed with multiple models with the doctors. (Figure 2E-F).

## Discussion

The modeled scenarios demonstrate a marked impact of staffing levels on MRSA acquisition rates. Consistently, over all models, adjusting nursing ratios resulted in fairly pronounced changes, while adding intensivists had a much smaller impact. Adding a second (or third) intensivist to the staff of an ICU never resulted in changes on par to decreasing the patient to nurse ratio. While adding a second physician to help the first dropped approximately 1 MRSA acquisition per 1000 person-years, adding a third had very slight changes from the second. This is most likely due to a physicians’ lower contact rate with patients in the first place. Adding a third cuts the contact rate to a minimal amount, especially in the finite tasks model (Table 1). This, of course, does not speak to other impacts, both positive and negative, in-patient care that an increase in intensivists might have^26^.

While the trends in the impact of staffing were consistent between the Infinite and Finite tasks models, both the absolute and relative values were markedly different, with the infinite task models having a much more linear effect that showed some evidence of levelling off at an (admittedly dire) 6:1 or 9:1 patient: nurse ratio, while the number of MRSA cases in the finite model continued to rise exponentially as the ratio increased. This difference comes down to the way the models are parameterized, and how nurse-to-patient contact interacts with the low level of largely transitory hand contamination present in both models^15^. In the infinite tasks model, the overall contact rate with a patient increases with each additional nurse, so the reduction in risk emerges from the dilution of any single nurse with contaminated hands into a larger overall pool of nurses. In contrast, in the finite task model, the addition of another nurse not only dilutes any contaminated nurses into a larger overall pool, but also reduces the number of times a contaminated nurse will contact a patient, owing to the smaller number of patient care tasks they are now responsible for.

## Conclusion

Our models show that, all else being equal, ICU staffing levels have a potentially dramatic increase on MRSA acquisition rates, and suggest that staffing should be considered a component of infection prevention – and that reduced levels of HAIs should be factored in when considering staff budgeting. These gains are largely seen in levels of nursing staff as compared to physicians. In both the Finite or Infinite models, there was no discernable difference between having two or three physicians in the ICU, and the addition of a single intensivist was modest. There is an opportunity for further research evaluating the economic costs and resulting benefits from these changes.

How one conceptualizes the workload of healthcare workers had a much more dramatic impact than any given staffing level on the modeled outcomes. While the infinite workload models had a steady increase in MRSA acquisitions as the nursing ratio in the ICU increased, the finite workload models showed a more exponential-like growth. One can imagine hospitals operating under both workloads. For example, a community hospital with a relatively stable catchment population might reasonably be modeled as having a finite workload within the ICU, whereas a busy critical access hospital (or that same community hospital during a public health emergency) might be more properly thought of as having an effectively infinite workload. For many hospitals, this may be reasonably assumed to vary from ward to ward.

It is then important to understand and act on the workload any given hospital or ward might be operating under when setting expectations for the impact of hiring additional staff. Incorrectly assuming an ICU has a finite workload has the potential to cause a severe overestimation of the impact of adding an additional nurse or changing ratios on infection rates.

This phenomenon is also important for the modeling of healthcare-associated infections. It highlights the importance of what seem like relatively benign and somewhat philosophical decisions as to how hospital workloads are modeled to the eventual outcomes of these models. It is essential, when considering model generalizability, to state the assumptions made about how workload and contact patterns within a hospital work, and to ensure these are appropriately tailored for the specific setting being modeled.

In conclusion, our models’ examination of ICU staffing’s impact on MRSA acquisition rates underscores the pivotal role that patient to nurse ratios play in infection prevention. The evidence suggests a potential for significant reduction in HAIs with optimized staffing, urging stakeholders to consider staffing as an integral component of infection control strategies. Notably, conceptualizing healthcare worker workload, rather than focusing solely on staffing ratios, significantly influenced modeled outcomes. Hospitals should tailor staffing decisions accordingly, recognizing the dynamic nature of their wards’ workload and adjusting accordingly. Importantly, they should consider the risk of over or underestimating the impact of additional staff on infection rates arises when workload assumptions are inaccurately defined. As healthcare models must inherently simplify complex scenarios, our study reinforces their value in capturing essential hospital dynamics. While our focus on nursing and physician staffing might exclude other healthcare workers, the deliberate simplifications enhance the clarity of our results, emphasizing the direct impact of staffing on infections transmitted by healthcare staff. Moving forward, recognizing the broader context of workload in a ward is crucial for refining strategies, setting expectations, and ensuring the generalizability of infection control interventions in dynamic healthcare settings.

## Data Availability

All data produced are available online at https://github.com/epimodels/StaffRatios

---

**Table S1.** Transitions and Equations for the Metapopulation Model of MRSA Acquisition.

| Process | Event | Transition | Equation |
| --- | --- | --- | --- |
| MRSA Acquisition & Transmission | Nurse Contaminated (Assigned Patient) | $N_{U,i}$ to $N_{C,i}$ | $\rho_N \sigma N_{Ui} \frac{P_{Ci}}{(P_{Ci} + P_{Ui})} \gamma ; \quad i = 1 \dots 18$ |
| | Nurse Contaminated (Unassigned Patient) | | $\rho_N \sigma N_{Ui} \frac{P_{Cj}}{(P_{Cj} + P_{Uj})} [(1 - \gamma) / 1 - i] ; \quad i = 1 \dots 18, j = 1 \dots 18, j \neq i$ |
| | Physician Contaminated | $D_U$ to $D_C$ | $\rho_D \sigma D_{Ui} \frac{\sum_{i=1}^x P_{Cj}}{\sum_{i=1}^x (P_{Cj} + P_{Uj})} ; \quad i = 1 \dots 3, j = 1 \dots 18, x = 1 \dots 18$ |
| | Patient Colonized (Assigned Nurse Contact) | $P_{U,i}$ to $P_{C,i}$ | $\rho_N \psi P_{Ui} \frac{N_{Ci}}{(N_{Ci} + N_{Ui})} \gamma ; \quad i = 1 \dots 18$ |
| | Patient Colonized (Unassigned Nurse Contact) | | $\rho_N \psi P_{Ui} \frac{N_{Cj}}{(N_{Cj} + N_{Uj})} [(1 - \gamma) / 1 - i] ; \quad i = 1 \dots 18, j = 1 \dots 18, j \neq i$ |
| | Patient Colonized (Physician Contact) | $P_{U,i}$ to $P_{C,i}$ | $\rho_D \psi P_{Ui} \frac{D_{Cj}}{(D_{Cj} + D_{Uj})} ; \quad i = 1 \dots 18, j = 1 \dots 3$ |
| MRSA Decolonization | Natural De-colonization | $P_{C,i}$ to $P_{U,i}$ | $\mu P_{Ci} ; \quad i = 1 \dots 18$ |
| Hand Hygiene and Decontamination | Nurse Hand Decontamination (Assigned Patient) | $N_{C,i}$ to $N_{U,i}$ | $\iota_N N_{Ci} ; \quad i = 1 \dots 18$ |
| | Physician Hand Decontamination | $D_{Ci}$ to $D_{Ui}$ | $\iota_D D_{Ci} ; \quad i = 1 \dots 3$ |
| | Nurse PPE Change (Assigned Patient) | $N_{C,i}$ to $N_{U,i}$ | $\tau_N N_{Ci} \frac{P_{Ci}}{(P_{Ci} + P_{Ui})} \gamma ; \quad i = 1 \dots 18$ |
| | Nurse PPE Change (Unassigned Patient) | | $\tau_N N_{Ci} \frac{P_{Cj}}{(P_{Cj} + P_{Uj})} [(1 - \gamma) / 5] ; \quad i = 1 \dots 18, j = 1 \dots 18, j \neq i$ |
| | Physician PPE Change | $D_{Ci}$ to $D_{Ui}$ | $\tau_D D_{Ci} \frac{\sum_{i=1}^x P_{Cj}}{\sum_{i=1}^x (P_{Cj} + P_{Uj})} ; \quad i = 1 \dots 3, j = 1 \dots 18, x = 1 \dots 18$ |
| Patient Admissions and Discharge | $P_{U,i}$ Discharge to $P_{U,i}$ Admission* | | $\theta v_U P_{Ui} ; \quad i = 1 \dots 18$ |
| | $P_{U,i}$ Discharge to $P_{C,i}$ Admission* | | $\theta v_C P_{Ui} ; \quad i = 1 \dots 18$ |
| | $P_{C,i}$ Discharge to $P_{U,i}$ Admission* | | $\theta v_U P_{Ci} ; \quad i = 1 \dots 18$ |
| | $P_{C,i}$ Discharge to $P_{C,i}$ Admission* | | $\theta v_C P_{Ci} ; \quad i = 1 \dots 18$ |

\* Note that patient discharge to patient admissions are not true “transitions” of a single individual, but rather the instantaneous replacement of a discharged patient with a newly admitted patient to maintain a steady population state.

## eMethods

Modeling Infectious Diseases in Healthcare Model Description Framework

## Purpose and Scope

### Purpose

The purpose of this model is the estimation of MRSA acquisitions based on staffing levels and approximate staff workloads found in the empirical literature.

### Scope

A single 18-bed intensive care unit in a U.S. academic medical center. This ICU is represented as a closed ICU, with no interaction with the rest of the hospital.

## Entities, state variables, and scales

### Entities

Patients, Nurses and Doctors. Interaction is defined entirely based on potentially contaminating/transmitting interactions. As a result, patients interact with nurses and doctors but not each other, while nurses and doctors both interact with patients but, again, not with each other. Nurses are specifically restricted to interacting with their assigned patient group 85% of the -me and then 15% with patients outside their assigned group.

### State Variables

Patients are classified as Uncolonized and Colonized. Patients are further segregated into six distinct groups, representing patients assigned to a particular nurse. Nurses and Doctors are represented as either Uncontaminated or Contaminated and are identified individually within the model.

### Scale

An 18-bed intensive care unit simulated for one year.

#### Initialization

In the initial state of the model (at time = 0), there are six uncontaminated nurses, one uncontaminated doctor, and six groups of three patients, all of whom are uncolonized. Further discussion of the effects of varying this initial state may be found in^1^. No burntiin period was used in the model as visual inspection suggested this initial state was relatively close to the stochastic equilibrium of the model.

## Process Overview and Scheduling

A full description of the processes of the model may be found in the supplement table, and for brevity are not presented here. The model is simulated using Gillespie’s Direct Method^2^, which selects the time the next event of any type occurs, and then randomly determines what type of event occurs based on their respective rates. As such, there is no overlying scheduling structure.

### Input Data

The model uses no external input data to represent processes in the model.

## Agent interactions and organism transmission

### Interactions

Interactions are event driven and concentrated on the interactions between healthcare workers (HCWs) and patients. At a model’s given rate per hour *ρN* for nurses and *ρD* for doctors, HCWs engage in a “direct care task”^3^ which involves touching the patient or their immediate surrounding environment.

This interaction prompts many other possible events in the model, including pathogen transmission (described below), HCW hand/body contamination, hand washing, and the donning/doffing of PPE by HCWs.

Patients do not interact with other patients directly – all patient-to-patient conflict is modeled as indirect interactions via shared and contaminated HCWs.

### Pathogen Transmission

Pathogen transmission is entirely indirect. A patient who is colonized (or has contaminated their environment) can contaminate a HCW they have come into contact with. If this HCW does not clear this contamination either by washing their hands or by removing contaminated PPE, there is a per-direct care task probability (ψ) that an uncolonized patient will be successfully colonized, representing a within-healthcare facility transmission event.

## Stochasticity

Due to the model’s implementation using Gillespie’s Direct Method^2^, the times events occur, and which event triggers at a given time are fully stochastic in the model. All other elements of the model, such as population size and parameter values, are deterministic.

### Submodels

This model has no submodels.

## Model verification, calibration and validation

### Verification

The model’s code was based on a previously published model^4^. All code used in the model was subject to code review, and several extreme value tests (setting particular parameter values to very high or very low values that should subsequently result in implausible results) were conducted.

### Calibration and Validation

The baseline model was calibrated to the ψ as in a previously published study and gave us a baseline average incidence of 8.3 MRSA acquisitions per 1000 person-years ^4^. The baseline parameter for “direct care tasks” were taken from Ballerman *et al*^3^ and used as the fixed number of tasks a HCW can do per hour for our finite models. This was then altered to the proportion of HCW in the ICU from the baseline for the infinite models.

